# The impact of changes in age-based eligibility criteria on seasonal influenza vaccine uptake in England between 2019 and 2024: A retrospective cohort study

**DOI:** 10.64898/2026.06.19.26356025

**Authors:** Anne M. Suffel, Jemma Walker, Eleanor V. H. Barry, Colin N.J. Campbell, Jamie Lopez Bernal, Suzanna L. R. McDonald, Helen I. McDonald, Sandra Mournier-Jack, Edward P. K. Parker

## Abstract

**Objectives:** To examine changes in seasonal influenza vaccine uptake among clinical risk groups over periods of differing age-based eligibility.

**Design:** Retrospective cohort study.

**Setting:** Individuals in England registered in the Clinical Practice Research Datalink Aurum.

**Participants:** Between 1,239,802 (2019/20) and 1,289,330 (2023/24) individuals aged 40-69 years in clinical risk groups.

**Interventions:** Natural experiment involving temporary expansion of age-based eligibility for influenza vaccination to include 50–64-year-olds from 2020/21 to 2022/23.

**Main outcome measures:** Influenza vaccine uptake from 1^st^ September to 28^th^ February, incidence rate ratio (IRR) of vaccine uptake across consecutive seasons within age groups, and the ratio of IRRs between age groups.

**Results:** Influenza vaccine uptake increased in all age groups in 2020/21 relative to 2019/20. The increase was larger in individuals aged 50-64 years (13.3%; IRR 1.50, 95% CI 1.50-1.51) compared with those aged 40-49 years (8.3%; IRR 1.35, 95% CI 1.34-1.35) and 65-69 years (6.8%; IRR 1.34, 95% CI 1.33-1.35). From 2020/21 to 2022/23, vaccine uptake decreased, with a more pronounced decline among those aged 40-49 years (−5.4%) compared with age-eligible groups (50-64 years: −3.0%; 65-69 years: −3.1%). The reversion of age eligibility in 2023/24 was associated with a larger decrease in uptake among those aged 50-64 years (−9.6% vs 2022/23; IRR 0.79, 95% CI: 0.79-0.79) compared with those aged 40-49 years (−4.9%; IRR 0.87, 95% CI: 0.87-0.88) and 65-69 years (−3.3%; IRR 0.97, 95% CI: 0.96-0.97). Patterns were broadly consistent across clinical risk groups.

**Conclusions:** The COVID-19 pandemic saw a general increase in seasonal influenza vaccine uptake in clinical risk groups. This increase was larger and more sustained in 50-64 year-olds who had also become eligible based on age. Our findings highlight the potential gains in vaccine coverage among clinical risk groups based on expanded age-based eligibility.

**Summary:** *What is already known on this topic:* Seasonal influenza presents a substantial public health burden. Individuals with underlying health conditions have a higher risk of morbidity and mortality from influenza and are eligible for the seasonal influenza vaccine. However, uptake among individuals in clinical risk groups in England remains suboptimal.

*What this study adds:* We examined whether changes in the age-eligibility threshold for the influenza vaccine from 65 years to 50 years during the COVID-19 pandemic in England (2020/21 to 2022/23) improved uptake among individuals at clinical risk of severe influenza. We found that uptake increased most and was more sustained in 50-64 year-olds who were eligible for vaccination by both age and clinical risk. Our findings highlight the potential gains in vaccine coverage among clinical risk groups based on expanded age-based eligibility.

## Introduction

Seasonal influenza presents a significant public health burden. There were 18,500 estimated annual deaths in 2017/18 due to influenza in England and Wales, and the virus acts as a significant driver of winter pressure on the NHS [1]. The risk of mortality from influenza is particularly pronounced in individuals with underlying health conditions such as immunosuppression, chronic liver disease, and chronic neurological disease [2].

The seasonal influenza vaccination programme in the UK aims to directly protect individuals at risk of severe influenza and reduce community transmission to further protect vulnerable individuals and reduce pressure on the NHS [3,4]. In 2019/20, vaccination was recommended for everyone aged 65 years or more, those aged six months to 65 years with a relevant underlying health condition or severe obesity (‘clinical risk groups’), close contacts of immunocompromised individuals, residents of long-stay residential care homes, carers, frontline health and social care workers, pregnant women, and all children aged 2-10 years [4]. After the onset of the COVID-19 pandemic, eligibility was temporarily widened to all adults over 50 years from 1^st^ December 2020 for the 2020/21 season [5] - prompted by concerns over the risk posed by influenza and SARS-CoV-2 co-circulation [6]. Expanded age eligibility was retained for the 2021/22 and 2022/23 seasons, before the age threshold reverted to 65 years in 2023/24.

The expansion and subsequent retraction of eligibility for seasonal influenza vaccination prompted by the COVID-19 pandemic provide natural experiments into the potential impact of expanded age-based eligibility on vaccine uptake in clinical risk groups.

## Methods

We conducted a retrospective cohort study using routinely collected primary care data from England between 2019 and 2024 to assess the impact of changes in age-based eligibility criteria on influenza vaccine uptake across different seasons. A RECORD statement can be found in the supplementary materials.

### Study population

Our study was conducted among individuals in England in the Clinical Practice Research Datalink (CPRD) Aurum primary care database (build: June 2024) [7]. The CPRD Aurum release contains information on 47,766,944 research quality patients [7] and is broadly representative of the English population based on age, sex, and ethnicity [8]. It holds information on demographics (sex, year of birth, region), clinical events (symptoms, diagnoses), primary care prescriptions, vaccination, and lifestyle information (e.g., smoking status).

We evaluated separate cohorts for each influenza season between 2019/20 and 2023/24. In each cohort, we included individuals with at least one underlying health condition that would make them eligible for a seasonal influenza vaccine (Table 1). All code lists were developed according to best practice guidelines [9], updating established code lists [10] using search terms aligned with codes from the PRIMIS specification used for routine reporting of vaccine coverage data for all GP practices in England [11]. The code to create the code lists can be found on our GitHub repository [12].

**Table 1:**
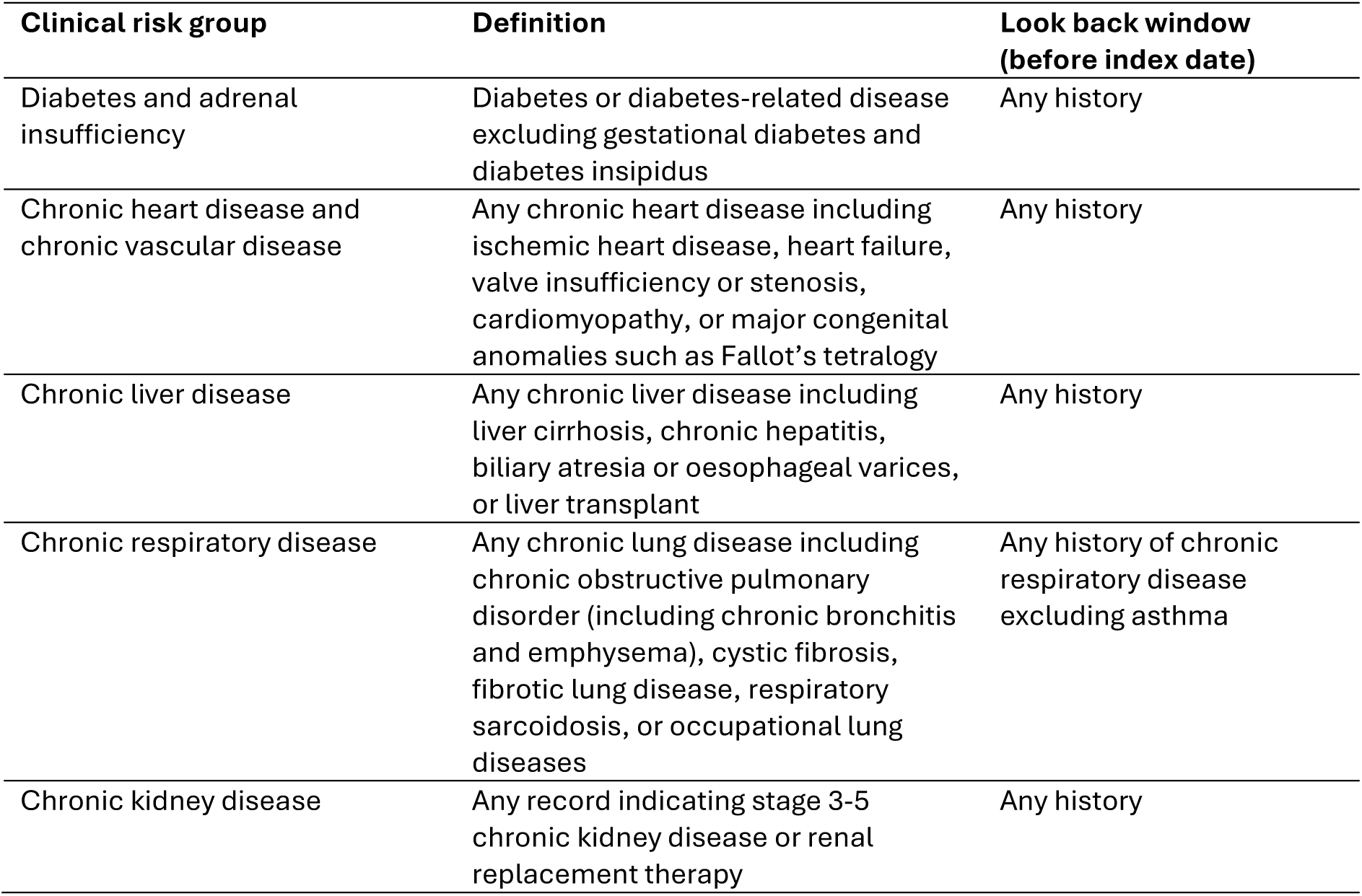
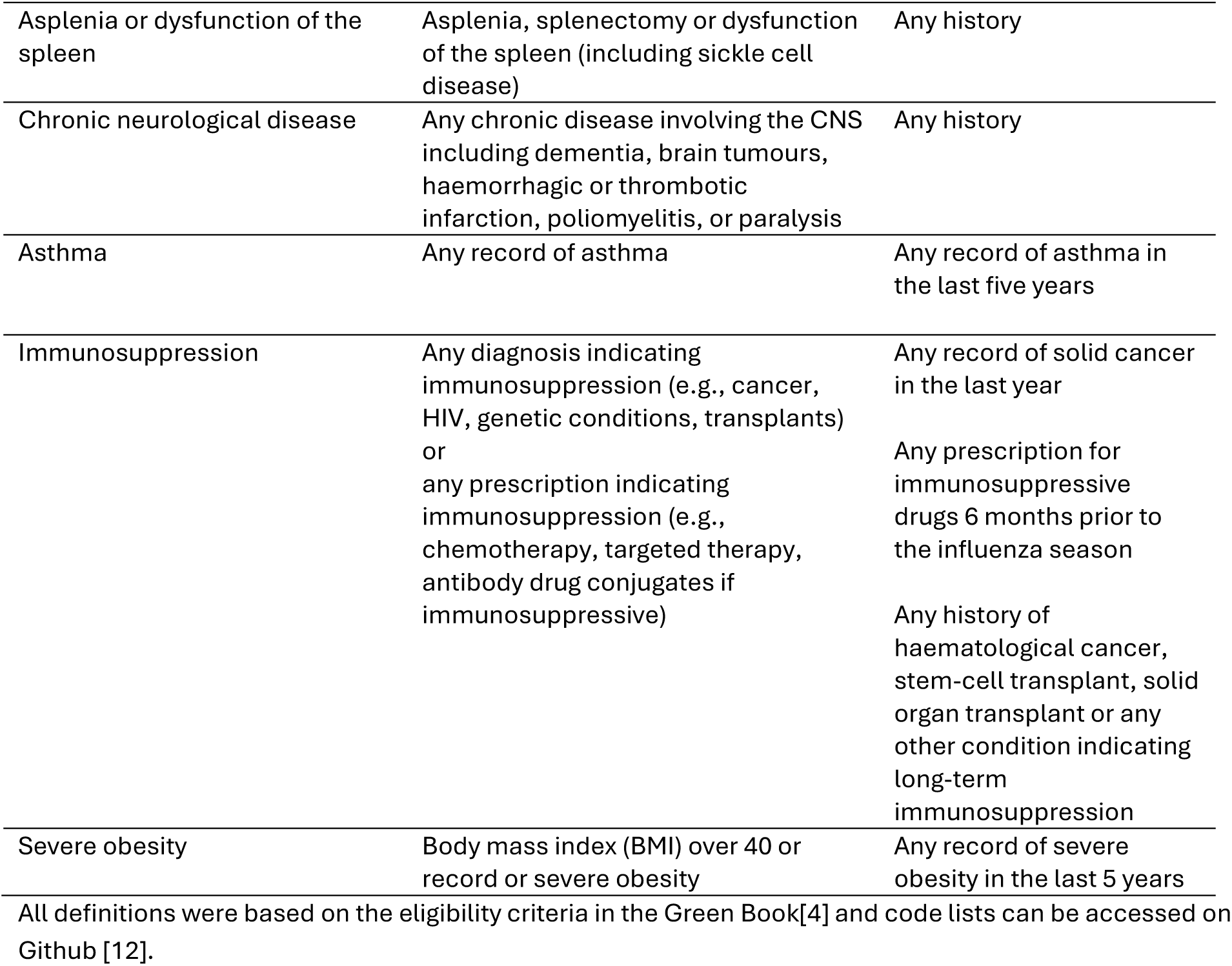
Definition of clinical risk groups.

| **Clinical risk group** | **Definition** | **Look back window**<br>**(before index date)** |
| --- | --- | --- |
| Diabetes and adrenal insufficiency | Diabetes or diabetes-related disease excluding gestational diabetes and diabetes insipidus | Any history |
| Chronic heart disease and chronic vascular disease | Any chronic heart disease including ischemic heart disease, heart failure, valve insufficiency or stenosis, cardiomyopathy, or major congenital anomalies such as Fallot's tetralogy | Any history |
| Chronic liver disease | Any chronic liver disease including liver cirrhosis, chronic hepatitis, biliary atresia or oesophageal varices, or liver transplant | Any history |
| Chronic respiratory disease | Any chronic lung disease including chronic obstructive pulmonary disorder (including chronic bronchitis and emphysema), cystic fibrosis, fibrotic lung disease, respiratory sarcoidosis, or occupational lung diseases | Any history of chronic respiratory disease excluding asthma |
| Chronic kidney disease | Any record indicating stage 3-5 chronic kidney disease or renal replacement therapy | Any history |
| Asplenia or dysfunction of the spleen | Asplenia, splenectomy or dysfunction of the spleen (including sickle cell disease) | Any history |
| Chronic neurological disease | Any chronic disease involving the CNS including dementia, brain tumours, haemorrhagic or thrombotic infarction, poliomyelitis, or paralysis | Any history |
| Asthma | Any record of asthma | Any record of asthma in the last five years |
| Immunosuppression | Any diagnosis indicating immunosuppression (e.g., cancer, HIV, genetic conditions, transplants) or any prescription indicating immunosuppression (e.g., chemotherapy, targeted therapy, antibody drug conjugates if immunosuppressive) | Any record of solid cancer in the last year<br>Any prescription for immunosuppressive drugs 6 months prior to the influenza season<br>Any history of haematological cancer, stem-cell transplant, solid organ transplant or any other condition indicating long-term immunosuppression |
| Severe obesity | Body mass index (BMI) over 40 or record of severe obesity | Any record of severe obesity in the last 5 years |

All definitions were based on the eligibility criteria in the Green Book[4] and code lists can be accessed on Github [12].

We included individuals between 40 and 69 years of age, thereby covering the age band affected by changes in age eligibility over the study period (50-64 years), as well as those below (40-49 years) and above (64-69 years) this age group. Follow-up began on 1^st^ September each season. Individuals were required to have at least one year of registration before the start of a given season and to meet CPRD quality-control standards at individual and practice level [13]. Individuals were excluded if they were ineligible for linkage to Office for National Statistics death registration and Hospital Episode Statistics. As our focus was on adults affected by the changes of age eligibility, we did not include pregnant women. Follow-up ended at the earliest of: vaccination, death (earliest date recorded in CPRD or ONS mortality data), the end of the influenza season (28^th^ February of the successive year), last collection date of the practice, or end of registration.

### Outcome

The receipt of a seasonal influenza vaccine was identified based on the combination of clinical and prescription codes adapting a validated algorithm (Supplementary Methods) [14]. Only codes recorded within each influenza season were used to determine vaccination status. Where there was more than one record of influenza vaccination, the first recorded date of administration was used.

### Covariates

Demographic covariates such as age and region were directly derived from the patient file in CPRD. Since CPRD provides only year of birth for adults to maintain anonymity, we assigned all participants a mid-year (1^st^ July) date of birth. Ethnicity categories (White, Black, South Asian, mixed, other) were identified based on clinical records using an established algorithm [15]. These broader categories were used to ensure positivity in the analysis. Index of multiple deprivation (IMD) in quintiles was derived based on small area data linkage [16]. We used practice-level IMD to supplement deprivation status for those without a record of individual-level deprivation.

### Statistical analysis

We described the uptake of seasonal influenza vaccine for each season overall and stratified by age group and clinical risk group.

To compare changes in influenza vaccine uptake across consecutive seasons, we used multivariate Poisson regression with log-offset for follow-up time to account for censoring. For each age band (40-49, 50-64, and 65-69 years), we calculated the incidence rate ratio (IRR) of influenza vaccine uptake for each season compared to the preceding season. We then compared the ratio of IRRs for individuals aged 50-64 years with those aged 40-49 years and (separately) those aged 65-69 years in 2020/21 relative to 2019/20 (expansion of age eligibility). We repeated the same analysis comparing these age groups in 2021/22 relative to 2020/21 (no change of age eligibility), 2022/23 relative to 2021/22 (no change of age eligibility), and 2023/24 relative to 2022/23 (reversion of age eligibility). Robust standard errors using the Hubert-White sandwich estimator were used to account for clustering by individual [17]. We then stratified the same analysis by underlying health condition. As ethnicity, sex, region, and socioeconomic status were not expected to change across successive years in the study population, we did not adjust for them in the regression model. A schematic of the overall study design is provided in Figure 1A.

**Figure 1.**
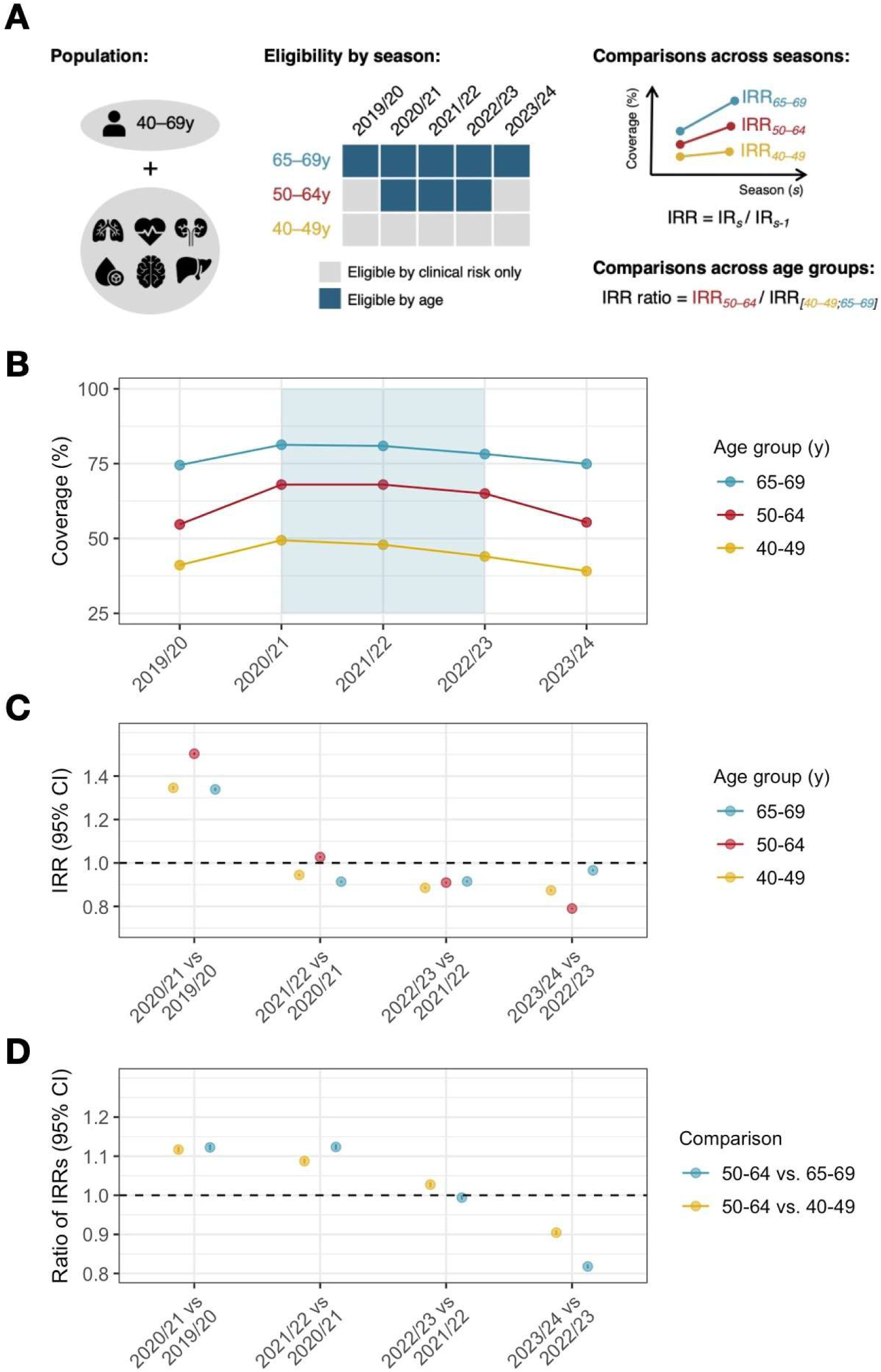
Impact of changes in age-based eligibility on influenza vaccine uptake among adults in clinical risk groups. (A) Study design. The study population for each season included individuals aged 40-69 years in at least one clinical risk group. Age-based eligibility was expanded to include 50-64 year-olds from 2020/21 to 2022/23. In 2020/21, 50-64 year-olds were eligible by age from 1^st^ December 2020. IRRs of influenza vaccine uptake were compared within age groups across consecutive seasons and the ratio of IRRs were compared between age groups. Clinical risk group icons obtained from Flaticon.com. (B) Vaccine uptake by season (1^st^ September to 28^th^ February of the consecutive year) stratified by age group. Seasons of expanded age-based eligibility are highlighted via blue shading. (C) IRRs comparing vaccine uptake between consecutive seasons within age groups. (D) Ratio of IRRs comparing changes in influenza vaccine uptake between age groups. CI, confidence interval; IR, incidence rate; IRR, incidence rate ratio; y, years.

### Sensitivity analysis

As age-based eligibility in 2020/21 was only expanded to include 50-64 year-olds on 1^st^ December 2020, we conducted a sensitivity analysis comparing influenza vaccine uptake in 2021/22 (the first season in which 50-64 year-olds were eligible by age from the outset of vaccine implementation) relative to 2019/20.

### Patient and public involvement

Our study was informed by a public involvement workshop in 2022 discussing opportunities to improve uptake of seasonal influenza vaccination among individuals in clinical risk groups.

## Results

We included between 1,239,802 (2019/20) and 1,289,330 (2023/24) individuals aged 40-69 years and in at least one clinical risk group (see Figure S1 for flow chart of cohort selection). Baseline characteristics were generally consistent across seasons overall (Table 2) and in individual age groups (Table S1), although there was a marked increase in the completeness of ethnicity recording over the study period. Individuals aged 50-64 years made up 55.7-56.8% of the overall cohort. Common clinical risk groups included asthma (e.g., 31.6% in 2023/24), diabetes (30.6% in 2023/24), chronic liver disease (14.2% in 2023/24), and chronic heart disease (13.7% in 2023/24).

**Table 2.** Baseline characteristics of the study population.

| **Variable** | **2019/20** | **2020/21** | **2021/22** | **2022/23** | **2023/24** |
| --- | --- | --- | --- | --- | --- |
| **N** | 1239802 | 1269044 | 1281065 | 1281738 | 1289330 |
| **Median (IQR) follow-up in days** | 94 (46-180) | 67 (35-180) | 66 (38-180) | 67 (39-180) | 70 (35-180) |
| **Sex** |  |  |  |  |  |
| Female | 614541 (49.6) | 629430 (49.6) | 636373 (49.7) | 637778 (49.8) | 643393 (49.9) |
| **Age group (years)** |  |  |  |  |  |
| 40-49 | 304763 (24.6) | 310106 (24.4) | 308555 (24.1) | 306130 (23.9) | 305743 (23.7) |
| 50-64 | 691035 (55.7) | 713419 (56.2) | 724802 (56.6) | 727566 (56.8) | 730671 (56.7) |
| 65-69 | 244004 (19.7) | 245519 (19.3) | 247708 (19.3) | 248042 (19.4) | 252916 (19.6) |
| **Region** |  |  |  |  |  |
| North East | 42911 (3.5) | 43793 (3.5) | 44267 (3.5) | 43713 (3.4) | 43012 (3.3) |
| North West | 246385 (19.9) | 253684 (20) | 255439 (19.9) | 263634 (20.6) | 266448 (20.7) |
| Yorkshire & The Humber | 40383 (3.3) | 39919 (3.1) | 39648 (3.1) | 30257 (2.4) | 28966 (2.2) |
| East Midlands | 26791 (2.2) | 25028 (2) | 23843 (1.9) | 18646 (1.5) | 18393 (1.4) |
| West Midlands | 208039 (16.8) | 213618 (16.8) | 216464 (16.9) | 221521 (17.3) | 222791 (17.3) |
| East of England | 49985 (4) | 49222 (3.9) | 50169 (3.9) | 52212 (4.1) | 50893 (3.9) |
| London | 227098 (18.3) | 236095 (18.6) | 242400 (18.9) | 250980 (19.6) | 257089 (19.9) |
| South East | 244554 (19.7) | 251742 (19.8) | 254388 (19.9) | 255312 (19.9) | 257340 (20) |
| South West | 153656 (12.4) | 155943 (12.3) | 154447 (12.1) | 145463 (11.3) | 144398 (11.2) |
| **Ethnicity** |  |  |  |  |  |
| White | 863339 (69.6) | 884755 (69.7) | 939033 (73.3) | 979380 (76.4) | 997945 (77.4) |
| Asian | 100219 (8.1) | 107445 (8.5) | 113145 (8.8) | 121132 (9.5) | 126699 (9.8) |
| Black | 55506 (4.5) | 59370 (4.7) | 63474 (5) | 68329 (5.3) | 71780 (5.6) |
| Other | 15835 (1.3) | 17552 (1.4) | 19112 (1.5) | 21414 (1.7) | 24836 (1.9) |
| Mixed | 13795 (1.1) | 14742 (1.2) | 15624 (1.2) | 17015 (1.3) | 17850 (1.4) |
| Unknown | 191108 (15.4) | 185180 (14.6) | 130677 (10.2) | 74468 (5.8) | 50220 (3.9) |
| **IMD quintile** |  |  |  |  |  |
| 1 (least deprived) | 231882 (18.7) | 236724 (18.7) | 237797 (18.6) | 235137 (18.3) | 234437 (18.2) |
| 2 | 236708 (19.1) | 240518 (19) | 241207 (18.8) | 240420 (18.8) | 241086 (18.7) |
| 3 | 237074 (19.1) | 240585 (19) | 242965 (19) | 241614 (18.9) | 244492 (19) |
| 4 | 262579 (21.2) | 270427 (21.3) | 274630 (21.4) | 276087 (21.5) | 279094 (21.6) |
| 5 (most deprived) | 271559 (21.9) | 280790 (22.1) | 284466 (22.2) | 288480 (22.5) | 290221 (22.5) |
| **Clinical risk groups** |  |  |  |  |  |
| Diabetes | 362994 (29.3) | 371551 (29.3) | 381807 (29.8) | 386821 (30.2) | 393927 (30.6) |
| Chronic heart disease | 175513 (14.2) | 178302 (14.1) | 178854 (14) | 177432 (13.8) | 177047 (13.7) |
| Chronic liver disease | 121443 (9.8) | 135102 (10.6) | 151112 (11.8) | 165946 (12.9) | 182574 (14.2) |
| Chronic respiratory diseases | 148958 (12) | 147945 (11.7) | 142820 (11.1) | 139123 (10.9) | 138659 (10.8) |
| Chronic kidney disease | 90655 (7.3) | 87858 (6.9) | 87537 (6.8) | 87874 (6.9) | 89471 (6.9) |
| Asplenia | 31171 (2.5) | 32233 (2.5) | 32655 (2.5) | 32802 (2.6) | 33172 (2.6) |
| Chronic neurological disease | 108641 (8.8) | 112319 (8.9) | 113158 (8.8) | 113310 (8.8) | 113772 (8.8) |
| Asthma | 407849 (32.9) | 419395 (33) | 418613 (32.7) | 410562 (32) | 407400 (31.6) |
| Immunosuppression | 147126 (11.9) | 148131 (11.7) | 150274 (11.7) | 153367 (12) | 152933 (11.9) |
| Severe obesity | 118165 (9.5) | 124072 (9.8) | 126844 (9.9) | 129653 (10.1) | 133652 (10.4) |

Data are N (%) unless otherwise stated. The study population for each season included individuals aged 40-69 years in at least one clinical risk group. The number of individuals censored unvaccinated before the end of follow-up was 28213 (2.3%) in 2019/20, 24382 (1.9%) in 2020/21, 26505 (2.1%) in 2021/22, 21354 (1.7%) in 2022/23, and 28912 (2.2%) in 2023/24. IMD, index of multiple deprivation; IQR, interquartile range.

### 2020/21 vs 2019/20: expansion of age eligibility

In 2019/20, influenza vaccine uptake in our cohort was 55.3% overall (41.1%, 54.7%, and 74.5% for individuals aged 40-49 years, 50-64 years, and 65-69 years, respectively; Figure 1B). In 2020/21, marking the first year of expanded age eligibility, vaccine uptake rose to 66.0% overall (a 10.7% absolute increase compared to 2019/20), with age-specific uptake of 49.4% at 40-49 years (8.3% increase), 68.0% at 50-64 years (13.3% increase), and 81.3% at 65-69 years (6.8% increase).

The IRR for influenza vaccination in 2020/21 relative to 2019/20 was 1.35 (95% CI: 1.34-1.35) for individuals aged 40-49 years, 1.50 (95% CI: 1.50-1.51) for those aged 50-64 years, and 1.34 (95% CI: 1.33-1.35) for those aged 65-69 years (Figure 1C). The ratio of IRRs was 1.12 (95% CI: 1.11-1.12) comparing individuals aged 50-64 years with those aged 40-49 years and 1.12 (95% 1.12-1.13) compared with those aged 65-69 years (Figure 1D).

Vaccine uptake varied across clinical risk groups (e.g., from 48.7% in 40–69-year-olds with chronic liver disease to 66.4% among those with diabetes in 2019/20). Changes in 2020/21 relative to 2019/20 were generally consistent across clinical risk groups, albeit with more pronounced effect sizes for individuals with immunosuppression, asthma, chronic liver disease, and chronic neurological disease relative to other clinical risk groups (Table S2, Figure 2, and Figure S2).

**Figure 2.**
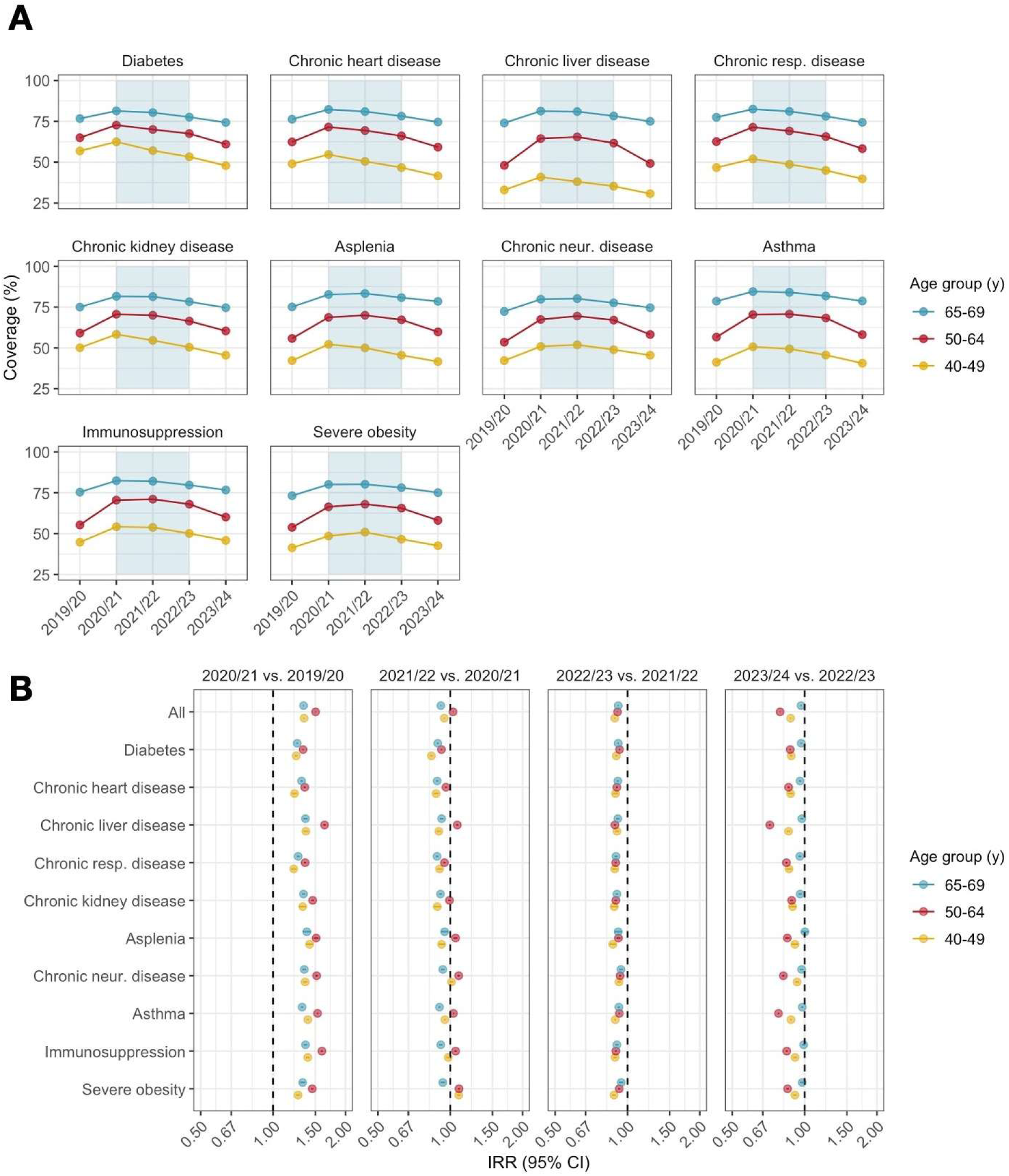
Impact of changes in age-based eligibility on influenza uptake across clinical risk groups. (A) Vaccine uptake by season (1^st^ September to 28^th^ February of the consecutive year) stratified by age and clinical risk group. Seasons of expanded age-based eligibility are highlighted via blue shading. In 2020/21, 50-64 year-olds were eligible by age from 1^st^ December 2020. (B) IRRs comparing vaccine uptake between consecutive seasons within age groups. See Figure S2 for plot of ratio of IRRs comparing changes in influenza vaccine uptake between age groups. IRR, incidence rate ratio; Neur., neurological; Resp., respiratory.

### 2021/22 and 2022/23: expanded age eligibility maintained

In 2021/22, vaccine uptake was similar to 2020/21 at 65.6% overall in those aged 40 to 69 years. Age-specific uptake was 47.9% among individuals aged 40-49 years (1.5% decrease compared to 2020/21; IRR 0.94 [95% CI 0.94-0.95]), 68.0% in those aged 50-64 years (0.0% change; IRR 1.03 [95% CI 1.02-1.03]), and 80.9% for those aged 65-69 years (0.4% decrease; IRR 0.91 [95% CI 0.91-0.92]; Figures 1B and 1C).

In 2022/23, vaccine uptake declined to 62.5% overall (44.0%, 65.0%, 78.2% for individuals aged 40-49 years, 50-64 years, and 65-69 years, respectively), with a more pronounced decline relative to 2021/22 among individuals aged 40-49 years (3.9% decrease; IRR 0.89 [95% CI 0.88-0.89) compared with those aged 50-64 years (3.0% decrease; IRR 0.91 [95% CI 0.91-0.91]) or 65-69 years (2.7% decrease; IRR 0.91 [95% CI 0.91-0.92).

Overall, comparing 2022/23 with 2020/21 (the last vs first seasons of expanded age eligibility), there was an absolute decrease in uptake of 3.5%, with a notably larger decline among individuals aged 40-49 years (5.4% decrease) compared with those above the age eligibility threshold (50-64 years: 3.0% decrease; 65-69 years: 3.1% decrease).

The decline in uptake from 2020/21 to 2022/23 among 40-49 years was generally consistent across clinical risk groups, with higher and more stable coverage observed in above risk groups above the age eligibility threshold (Table S2 Figure 2, and Figure S2).

### 2023/24 vs 2022/23: reversion of age eligibility

In 2023/24, marking the reversion of expanded age eligibility, influenza vaccine uptake declined to 55.4% overall (a 7.1% absolute decrease compared to 2022/23), with age-specific uptake of 39.1% at 40-49 years (4.9% decrease), 55.4% at 50-64 years (9.6% decrease), and 74.9% at 65-69 years (3.3% decrease; Figure 1B).

The IRR for influenza vaccination in 2023/24 relative to 2022/23 was 0.87 (95% CI: 0.87-0.88) for individuals aged 40-49 years, 0.79 (95% CI: 0.79-0.79) for those aged 50-64 years, and 0.97 (95% CI: 0.96-0.97) for those aged 65-69 years (Figure 1C). The ratio of IRRs was 0.90 (95% CI: 0.90-0.91) comparing individuals aged 50-64 years with those aged 40-49 years and 0.82 (95% CI: 0.81-0.82) compared with those aged 65-69 years (Figure 1C).

Across clinical risk groups, absolute declines in vaccine coverage among individuals aged 50-64 years (no longer age-eligible for vaccination) ranged from 6.0% in people with chronic kidney disease to 12.5% among those with chronic liver disease (Figure 2). IRR ratios comparing 50-64 year-olds with 65-69 year-olds were consistently below 1, indicating a more pronounced decrease in uptake in 2023/24 among individuals no longer eligible for vaccination based on age (Figure S2). The effect size was larger for individuals with chronic liver disease, mirroring the pattern observed from 2019/20 to 2020/21. When comparing 50-64 year-olds with 40-49 year-olds, patterns of vaccine uptake varied across clinical risk, with some groups (e.g., diabetes, chronic heart disease, and chronic kidney disease) exhibiting IRR ratios of approximately 1 (indicating parallel declines in coverage across age groups). By contrast, IRR ratios and 95% CIs were below 1 (indicating a larger decline in 50-64 year-olds) for individuals with severe obesity, chronic liver disease, immunosuppression, asthma, chronic neurological disease, and asplenia (Figure S2).

### 2023/24 vs 2019/20: vaccine coverage after vs before temporary change of age eligibility

Compared with 2019/20 (the season preceding the temporary expansion of age-based eligibility), we observed a slight overall increase in coverage in individuals aged 50-64 years (0.7%) and 65-69 years (0.4%), in contrast to a decline among those aged 40-49 years (−2.0%; Figure 3). Patterns differed substantially across clinical risk groups – coverage decreased in 2023/24 relative to 2019/20 across all age groups in individuals with diabetes, chronic heart disease, and chronic respiratory disease, but increased across all age groups in individuals with chronic neurological disease, immunosuppression, and severe obesity.

**Figure 3.**
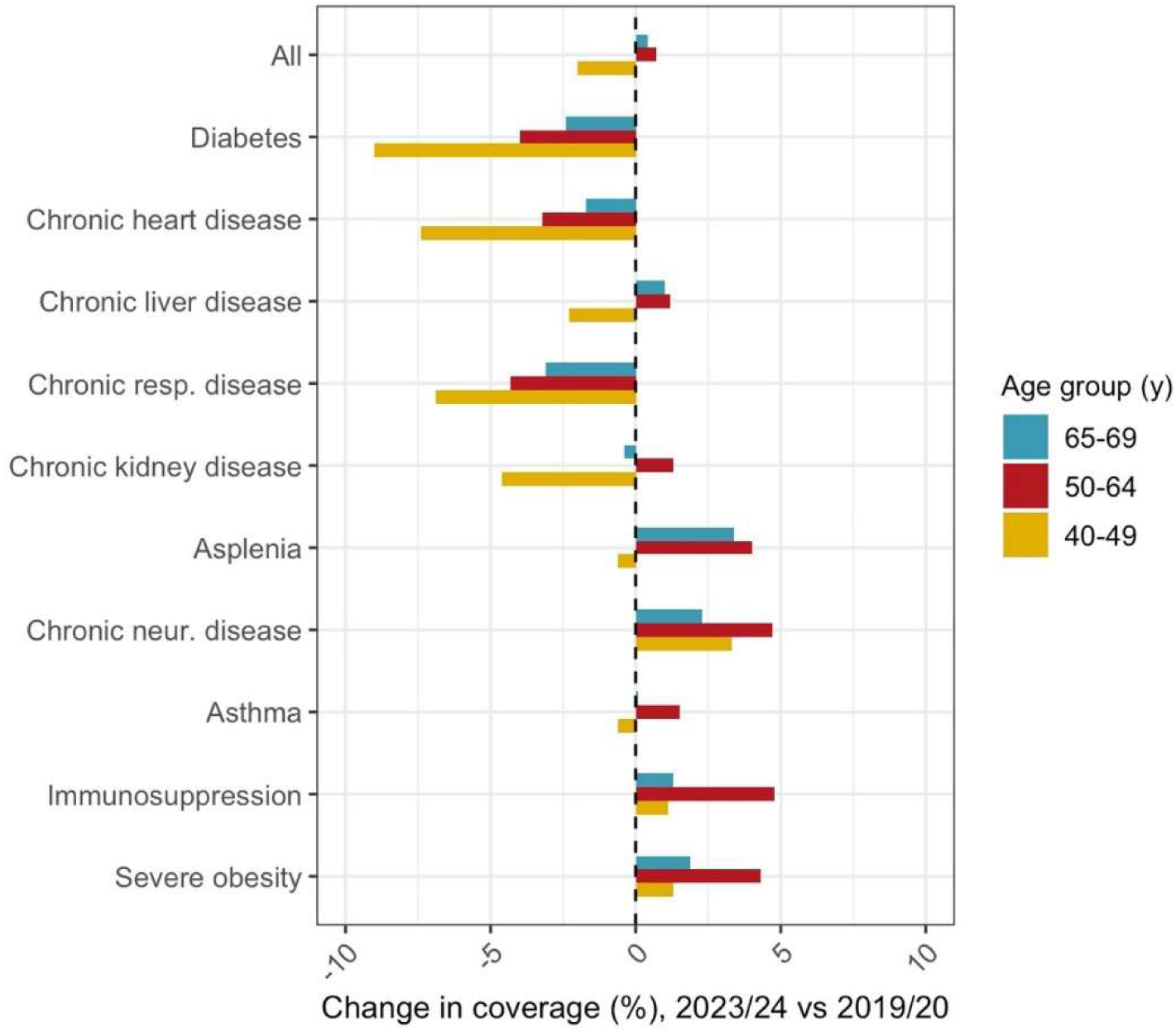
Changes in vaccine uptake in 2023/24 relative to 2019/20 among individuals aged 40-69 years in at least one clinical risk group. Estimates are presented overall and by clinical risk group. y, years.

### Sensitivity analysis

When comparing 2021/22 (the first season in which 50-64 year-olds were eligible by age for the entire influenza season) with 2019/20, effect sizes were larger than those observed in the primary analysis of expanded age eligibility (2020/21 vs 2019/20). The ratio of IRRs was 1.21 (95% CI: 1.20-1.22) comparing individuals aged 50-64 years with those aged 40-49 years and 1.26 (95% CI: 1.25-1.27) relative to those aged 65-69 years (Table S2).

## Discussion

The first autumn and winter of the COVID-19 pandemic saw a marked rise in seasonal influenza vaccine uptake among individuals in clinical risk groups in England. Compared with adjacent age groups, this increase in uptake was higher in 50-64 year-olds who were temporarily age-eligible for vaccination from 2020/21 to 2022/23. The subsequent reversion of age eligibility in 2023/24 was associated with a decline in coverage relative to 2022/23 by 9.6% among 50-64 year-olds – significantly larger than the decline in coverage seen in other age groups. Trends were consistent across clinical risk groups, albeit with greater gains (and subsequent falls) in coverage among 50-64 year-olds with chronic liver disease relative to other risk groups.

### Findings in context

A key challenge in interpreting our findings is the general impact of COVID-19 on health-seeking behaviour and healthcare access during the study period. This was evident through the increases in influenza vaccine coverage across age groups in 2020/21, including individuals aged 40-49 and 65-69 years who were unaffected by changes in eligibility criteria relative to 2019/20. The reversion of eligibility in 2023/24 – several years after the volatile early stages of the pandemic – may offer a clearer indication of the potential impact of changes in age-based eligibility in a typical influenza season. England has adopted a coverage target of 75% among adults over the age of 65 years, aligned with WHO recommendations for 2010 [18]. For clinical risk groups under the age of 65 years, 55% has been adopted as an interim coverage target, with 75% as a longer-term target [19]. Among 50-64 year-olds in our study, coverage was 55% in seasons in which individuals were eligible based only on clinical risk (2019/20 and 2023/24) but increased to 65-68% during the temporary expansion of age eligibility (with 68-71% coverage among 50-64 year-olds with immunosuppression).

It remains unclear why some clinical risk groups differed in their level of vaccine coverage in 2023/24 compared to 2019/20. Some groups may have benefited from increased communication related to their clinical risk as well as habituation of vaccination during the period of expanded age-based eligibility. For other groups, the perception of clinical risk may have been normalised and healthcare contact patterns may have changed in 2023/24 relative to 2019/20. Notwithstanding its pandemic context, our study highlights the potential of improving coverage in the most vulnerable groups through expanded age-based eligibility.

Qualitative research has shown that vaccine uptake in individuals in clinical risk groups often depends on the perceived risk of influenza infection as well as the relevance of the vaccine for their own condition [20]. Simple age-based criteria for defining vaccine eligibility may increase uptake as eligibility does not rely on clinical judgement (of either the affected individual or their healthcare provider) on whether a person falls within the clinical risk groups eligible for vaccination. Widening eligibility by age may also raise awareness of the programme through broader eligibility messaging and simplify access to it.

A study from the UK assessing seasonal and pandemic influenza vaccine uptake among clinical risk groups in 2009/10 found higher uptake of the respective vaccine if individuals were in age groups eligible for universal vaccination compared with age groups in which only clinical risk groups were eligible [21]. A Spanish study assessed the impact of the local expansion in age eligibility for the influenza vaccine in Madrid to cover individuals aged 60 years and above from 2005 (in contrast to an eligibility threshold of 65 years in the rest of the country), showing a significant increase in vaccine uptake among 60-64 year-olds with clinical risk conditions two years after the policy change [22].

There are currently an estimated 11.1 million individuals in the UK [23] over the age of 65 (18.6% of the population) who are eligible by age for annual influenza vaccination. There are a further 11.6 million 50-64 year-olds (19.5% of the population), of whom approximately 23.3% are eligible for influenza vaccination based on clinical risk [23,24]. In addition to promoting higher uptake in clinical risk groups, expanded age-based eligibility would also offer direct benefits to individuals without underlying health conditions, and indirect benefits to those they contact. In particular, 50-64 year-olds have a 3-fold increased risk of hospitalisation and a 9-fold increased risk of mortality due to influenza in comparison to those aged 18-49 years [25]. This may be caused by both a decline of immune competence in adults aged 50 and older, and an increasing presence of underlying health conditions (both diagnosed and undiagnosed) in adults over 50.

Multiple studies have explored the cost-effectiveness of extending age-based eligibility for seasonal influenza vaccination [26–28]. Notably, Baguelin et al [28] modelled multiple potential age-based extensions to the seasonal influenza vaccine programme in England and Wales, finding high uncertainty in the cost-effectiveness of extensions beyond 2-16 year-olds. In contrast, two other studies based in the USA and France found universal seasonal influenza vaccination for 50-64 year-olds to be cost-effective [29,30]. A general feature of economic modelling studies is that base scenarios fix vaccine coverage in risk groups and older adults (i.e., those with existing eligibility), while modelling various coverage scenarios in low-risk individuals. Our study suggests that future cost-effectiveness studies should incorporate increased coverage among high-risk individuals as an expected consequence of any age-based extension in eligibility.

### Strengths and limitations of the study

Strengths of our study include the scale and representativeness of CPRD Aurum, which covers 25% of the English population [8]. Using primary care data, it was possible to assess uptake across a comprehensive range of clinical risk groups eligible for seasonal influenza vaccination. The study cohorts in every influenza season were of comparable demographic characteristics, and we accounted for individual-level clustering through the use of robust standard errors. Using a ratio-of-ratios approach, we could evaluate changes in influenza vaccine uptake among 50-64 year-olds across seasons in the context of changing healthcare utilisation patterns in adjacent age groups that were not impacted by eligibility changes.

The main limitation of the study is its pandemic context, which makes it challenging to disentangle the influence of expanded age eligibility from altered risk-perception and communication during the ongoing pandemic. Influenza vaccines are also offered in pharmacies and other healthcare settings (e.g., certain outpatient facilities) [31]. The ascertainment from GP systems of vaccinations given in pharmacies differs by age, disease severity, and over time [32]. The clinical risk group definitions in the Green Book [4] provide examples of health conditions but also place an emphasis on clinical judgement regarding who should be invited for seasonal influenza vaccination. Our definitions of clinical risk groups in this study may not fully align with the population receiving invitations for vaccination. If this misalignment was greater for certain clinical risk groups (e.g., chronic liver disease), a larger increase in uptake would be expected during the period of unambiguous age-based eligibility as some individuals may not have received an invitation for the influenza vaccine when eligible based on clinical risk condition alone (i.e., before 2020/21).

## Conclusions

The UK’s seasonal influenza vaccination programme currently falls short of interim and long-term coverage targets that had previously been set for individuals in clinical risk groups under the age of 65 years – a group at high clinical risk of severe morbidity and mortality from influenza. Our study suggests that expansion of age-based eligibility – if economically and programmatically viable – can offer a useful approach towards simplifying vaccination recommendations and thereby improving uptake in high-risk populations.

## Contributors

Conceptualisation: AMS, SLRM, HIM, SM-J, EPKP; Methodology: AMS, EVHB, EPKP; Formal analysis: AMS; Writing–original draft: AMS; Writing–review & editing: AMS, JW, EVHB, CNJC, JLB, SLRM, HIM, SM-J, EPKP; Visualisation: AMS, EPKP; Project administration: AMS, EPKP. All authors had final responsibility for the decision to submit for publication. The corresponding author attests that all listed authors meet authorship criteria and that no others meeting the criteria have been omitted.

## Ethics statement

We received data governance approval from CPRD (protocol number 22_001750) and ethical approval from the London School of Hygiene and Tropical Medicine’s research ethics committee (reference number 27358).

## Data sharing

This study uses data from the Clinical Practice Research Datalink (CPRD) which does not allow sharing of patient-level data. The specification for the CPRD data set used in this study is available at: https://www.cprd.com/doi/cprd-aurum-june-2024-dataset. Analysis code and code lists are shared via a Github repository available at: https://github.com/Eyedeet/Influenza_uptake_England/.

## Acknowledgements

This study is based on data from the CPRD obtained under licence from the UK Medicines & Healthcare products Regulatory Agency. The data is provided by patients and collected by the NHS as part of their care and support.

