## Supplementary material for "The impact of changes in age-based eligibility criteria on seasonal influenza vaccine uptake in England between 2019 and 2024: A retrospective cohort study"

**Contents:**

P2: Supplementary Methods
P4: Supplementary Figures S1–S2
P6: RECORD checklist

**Supplementary Methods**

*Algorithm to ascertain influenza vaccination*

1. Classification of vaccination codes

We compiled all codes used in the drug issue file as codes indicating that any influenza vaccine product was given. Codes from the observation file were classified into: (i) codes indicating that a vaccine was administered; (ii) codes indicating that a vaccine was declined or an appointment not attended; and (iii) neutral codes mentioning the influenza vaccine. Codes of adverse reactions were not used as the timing of the vaccine could not be determined. Examples of different types of codes to capture influenza vaccines are provided in the table below:

| **Product code** | | **Administration code** | | **Decline code** | | **Neutral code** | |
|---|---|---|---|---|---|---|---|
| prodcodeid | Term | medcodeid | Term | medcodeid | Term | medcodeid | Term |
| 1043841000033118 | Pasteur Merieux Inactivated Influenza Vaccine 0.5 ml | 1193495100006116 | Administration of second non adjuvanted trivalent (TIV) inactivated seasonal influenza vaccination | 1727791000006112 | Influenza A virus subtype H1N1 vaccination declined | 6569221000006119 | Influenza vaccine |

Some codes indicated a delayed vaccination (e.g., "Has influenza vaccination at hospital; medcodeid: 816791000006117). These were treated as administration codes but interpreted differently once the date of the vaccine was determined (see below).

2. Identifying vaccination events

For each season, we counted the number of influenza vaccine records per day per patient. If there was more than one record per day, we evaluated the combination of influenza vaccine records to determine whether a patient was likely to have received an influenza vaccine. The interpretation of code type combinations is presented in the table below:

| **Combination of code types** | **Interpretation** |
|---|---|
| Neutral + administered | Vaccinated |
| Neutral + declined | Unvaccinated |
| Neutral + product | Vaccinated |
| Administered + product | Vaccinated |
| Administered + declined | Conflict |
| Product + declined | Conflict |
| Product + declined + Administered | Conflict |
| Neutral only | Vaccinated |
| Administered only | Vaccinated |
| Product only | Vaccinated |
| Declined only | Unvaccinated |

Code combinations interpreted as 'Conflict' or 'Unvaccinated' were then dropped. This ensured that for the assessment of the vaccination date, individuals who previously declined the vaccine would still show a record if they received the vaccine at a later date within a given season.

3. Determination of vaccination date

The first day during follow-up with codes evaluated as 'Vaccinated' (as defined above) was assigned as the day of vaccination. If an individual did not have any day with vaccination records evaluated as 'Vaccinated', they were classified as unvaccinated for a given season.

The following table illustrates the number of codes for the season 2019/20:

| | N overall* | N individuals with type of code* |
|---|---|---|
| *Limiting codes to codes within the influenza season* | | |
| Number of medcodes related to vaccination from the observation files | 1,442,979 | 1,202,926 |
| Number of product codes related to influenza vaccine | 197,379 | 485,332 |
| *Removing adverse events* | | |
| Number of medcodes related to vaccination from the observation files after removing adverse events | 1,427,407 | 836,507 |
| *Vaccination events after evaluation code combinations on the same day* | | |
| Vaccinated | 654,377 | – |
| (Vaccinated with delay) | 111,337 | – |
| Vaccination declined | 170,793 | – |
| Conflict codes | 846 | – |
| *Dropping conflict codes and unvaccinated codes* | | |
| Vaccination events | 685,231 | 685,073 |

* Some individuals receive multiple codes related to vaccination, leading to a difference between the absolute number of vaccination events and individuals with vaccination codes. All individuals with more than one vaccination event had either multiple records for a successful vaccination or a code with delayed vaccination. If there a multiple valid vaccination events, the earliest date was chosen.

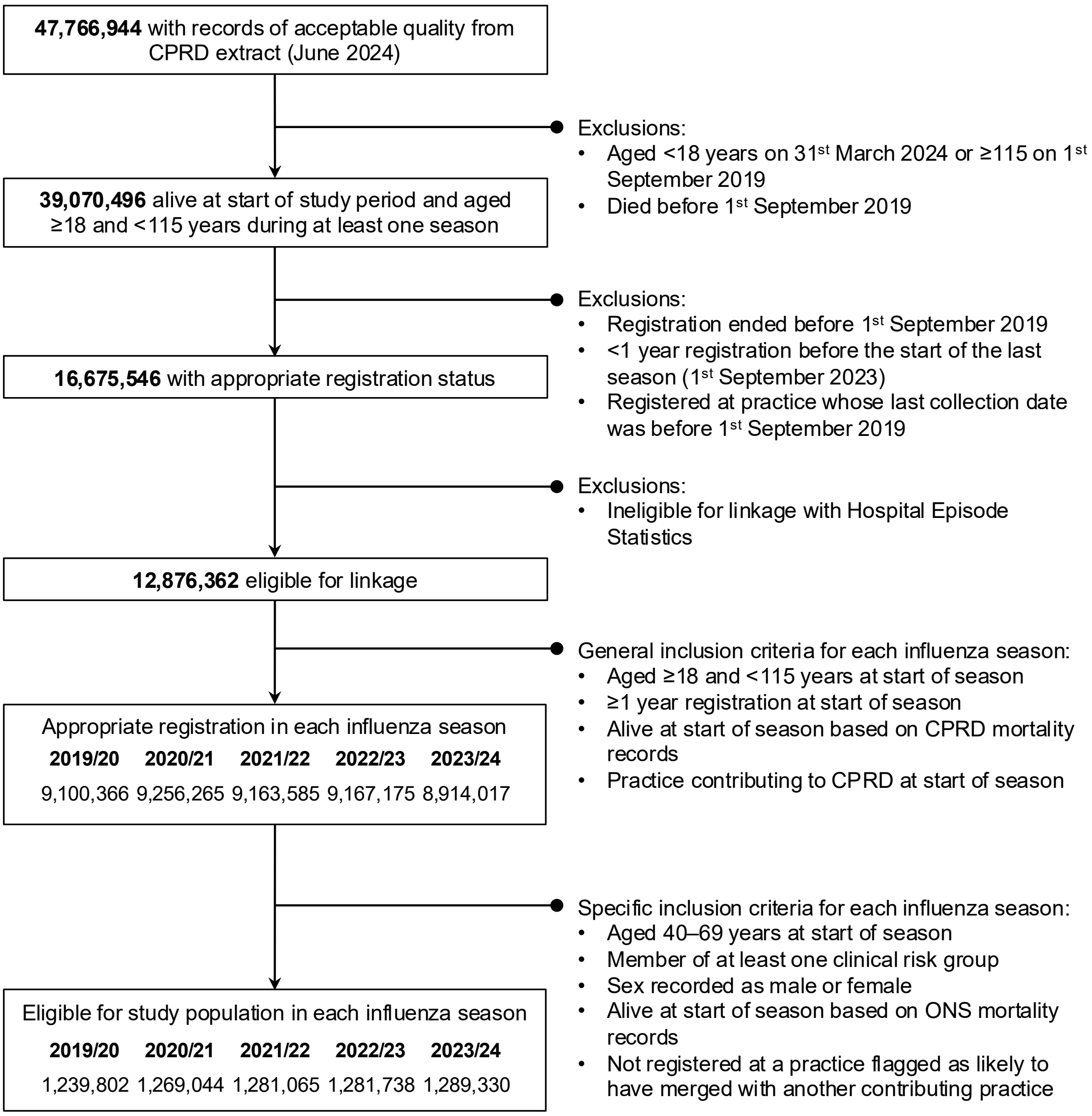


**Supplementary Figure S1: Flow chart of study population selection.** General inclusion criteria, including eligibility for linkage with Hospital Episode Statistics, were applied during cohort selection to support separate objectives within a broader study of factors associated with influenza vaccine uptake. CPRD, Clinical Practice Research Datalink; ONS, Office for National Statistics.

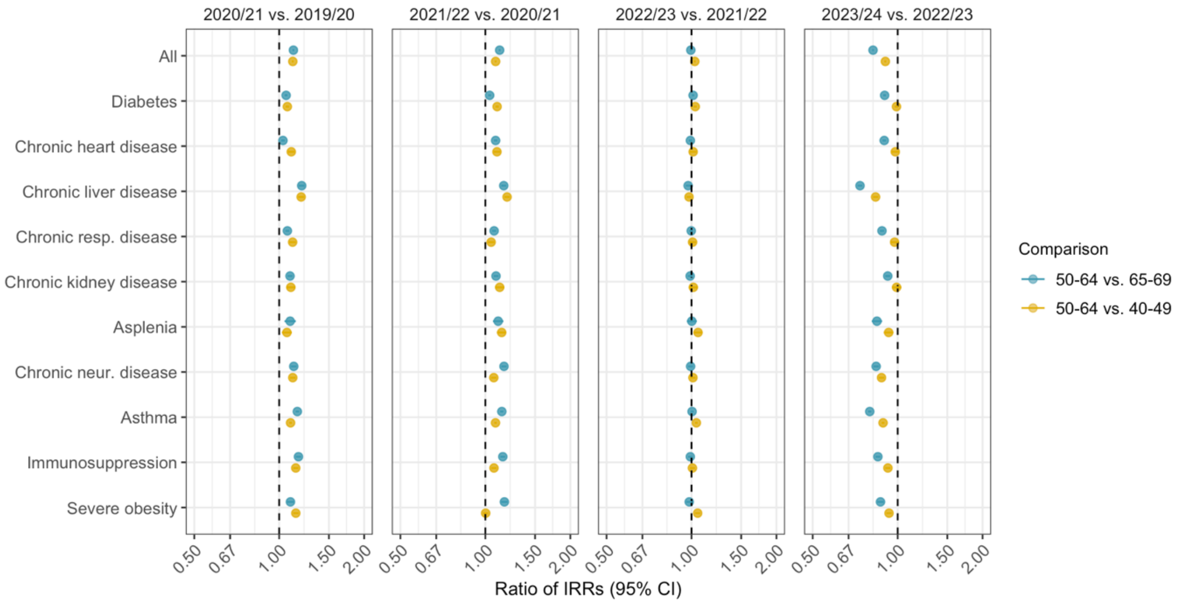


**Supplementary Figure S2: Ratio of incidence rate ratios by clinical risk group.** CI, confidence interval; IRR, incidence rate ratio.

**The RECORD statement – checklist of items, extended from the STROBE statement, that should be reported in observational studies using routinely collected health data.**

| | Item No. | STROBE items | Location in manuscript where items are reported | RECORD items | Location in manuscript where items are reported |
|---|---|---|---|---|---|
| **Title and abstract** | | | | | |
| | 1 | (a) Indicate the study's design with a commonly used term in the title or the abstract (b) Provide in the abstract an informative and balanced summary of what was done and what was found | Title, Abstract | RECORD 1.1: The type of data used should be specified in the title or abstract. When possible, the name of the databases used should be included.<br><br>RECORD 1.2: If applicable, the geographic region and timeframe within which the study took place should be reported in the title or abstract.<br><br>RECORD 1.3: If linkage between databases was conducted for the study, this should be clearly stated in the title or abstract. | Abstract<br><br>Title, Abstract<br><br>Linkage to secondary care, mortality, and small area data described in Methods (established linkages) |
| **Introduction** | | | | | |
| Background rationale | 2 | Explain the scientific background and rationale for the investigation being reported | Background | | |
| Objectives | 3 | State specific objectives, including any prespecified hypotheses | Background | | |
| **Methods** | | | | | |
| Study Design | 4 | Present key elements of study design early in the paper | Methods, Figure 1 | | |
| Setting | 5 | Describe the setting, locations, and relevant dates, including periods of recruitment, exposure, follow-up, and data collection | Methods ('Study population' and 'Statistical analysis' subheadings), Table 1, Supplementary methods (for influenza vaccination status) | | |
| Participants | 6 | *(a) Cohort study* - Give the eligibility criteria, and the sources and methods of selection of participants. Describe methods of follow-up<br>*Case-control study* - Give the eligibility criteria, and the sources and methods of case ascertainment and control selection. Give the rationale for the choice of cases and controls<br>*Cross-sectional study* - Give the eligibility criteria, and the sources and methods of selection of participants<br><br>*(b) Cohort study* - For matched studies, give matching criteria and number of exposed and unexposed | Methods ('Study population' and 'Statistical analysis' subheadings)<br><br>N/A | RECORD 6.1: The methods of study population selection (such as codes or algorithms used to identify subjects) should be listed in detail. If this is not possible, an explanation should be provided.<br><br>RECORD 6.2: Any validation studies of the codes or algorithms used to select the population should be referenced. If validation was conducted for this study and not published elsewhere, detailed methods and results should be provided.<br><br>RECORD 6.3: If the study involved linkage of databases, consider use of a flow diagram or other graphical display to demonstrate the data linkage | Methods, Supplementary Methods, Github repository<br><br>Validation of algorithm to define vaccination status referenced (Suffel et al, 2024; DOI 0.1002/pds.5848)<br><br>Linkage to secondary care. mortality, and small area data described in Methods (established linkages) |

| | | | | | |
|---|---|---|---|---|---|
| | | *Case-control study* - For matched studies, give matching criteria and the number of controls per case | | process, including the number of individuals with linked data at each stage. | |
| Variables | 7 | Clearly define all outcomes, exposures, predictors, potential confounders, and effect modifiers. Give diagnostic criteria, if applicable. | Methods ('Outcome', 'Covariates', and 'Statistical analysis' subheadings) | RECORD 7.1: A complete list of codes and algorithms used to classify exposures, outcomes, confounders, and effect modifiers should be provided. If these cannot be reported, an explanation should be provided. | Table 1, Supplementary methods (for influenza vaccination status), Github repository |
| Data sources/ measurement | 8 | For each variable of interest, give sources of data and details of methods of assessment (measurement).<br>Describe comparability of assessment methods if there is more than one group | Table 1, Github repository<br><br>N/A | | |
| Bias | 9 | Describe any efforts to address potential sources of bias | Methods ('Statistical analysis' subheading) | | |
| Study size | 10 | Explain how the study size was arrived at | Supplementary Figure 1 | | |
| Quantitative variables | 11 | Explain how quantitative variables were handled in the analyses. If applicable, describe which groupings were chosen, and why | Methods ('Covariates' subheading) | | |
| Statistical methods | 12 | (a) Describe all statistical methods, including those used to control for confounding<br>(b) Describe any methods used to examine subgroups and interactions<br>(c) Explain how missing data were addressed<br>(d) *Cohort study* - If applicable, explain how loss to follow-up was addressed<br>*Case-control study* - If applicable, explain how matching of cases and controls was addressed<br>*Cross-sectional study* - If applicable, describe analytical methods taking account of sampling strategy<br>(e) Describe any sensitivity analyses | Methods ('Statistical analysis' and 'Sensitivity analysis' subheadings) | | |
| Data access and cleaning methods | | .. | | RECORD 12.1: Authors should describe the extent to which the investigators had access to the database population used to create the study population.<br><br>RECORD 12.2: Authors should provide information on the data cleaning methods used in the study. | Data sharing<br><br>Table 1, Github repository |
| Linkage | | .. | | RECORD 12.3: State whether the study included person-level, institutional-level, or other data linkage across two or more databases. The methods of linkage and methods of linkage quality evaluation should be provided. | Methods ('Study population' and 'Covariates' subheadings), Supplementary Figure 1 |
| **Results** | | | | | |

| | | | | | |
|---|---|---|---|---|---|
| Participants | 13 | (a) Report the numbers of individuals at each stage of the study (*e.g.*, numbers potentially eligible, examined for eligibility, confirmed eligible, included in the study, completing follow-up, and analysed)<br>(b) Give reasons for non-participation at each stage.<br>(c) Consider use of a flow diagram | Supplementary Figure 1, Table 1, Results | RECORD 13.1: Describe in detail the selection of the persons included in the study (*i.e.*, study population selection) including filtering based on data quality, data availability and linkage. The selection of included persons can be described in the text and/or by means of the study flow diagram. | Supplementary Figure 1 |
| Descriptive data | 14 | (a) Give characteristics of study participants (*e.g.*, demographic, clinical, social) and information on exposures and potential confounders<br>(b) Indicate the number of participants with missing data for each variable of interest<br>(c) *Cohort study* - summarise follow-up time (*e.g.*, average and total amount) | Results, Table 1, Table S2 | | |
| Outcome data | 15 | *Cohort study* - Report numbers of outcome events or summary measures over time<br>*Case-control study* - Report numbers in each exposure category, or summary measures of exposure<br>*Cross-sectional study* - Report numbers of outcome events or summary measures | Results, Table S2 | | |
| Main results | 16 | (a) Give unadjusted estimates and, if applicable, confounder-adjusted estimates and their precision (e.g., 95% confidence interval). Make clear which confounders were adjusted for and why they were included<br>(b) Report category boundaries when continuous variables were categorized<br>(c) If relevant, consider translating estimates of relative risk into absolute risk for a meaningful time period | Table S2 | | |
| Other analyses | 17 | Report other analyses done—e.g., analyses of subgroups and interactions, and sensitivity analyses | Results ('Sensitivity analysis' subheading), Table S2 | | |
| **Discussion** | | | | | |
| Key results | 18 | Summarise key results with reference to study objectives | Discussion (first paragraph) | | |
| Limitations | 19 | Discuss limitations of the study, taking into account sources of potential bias or imprecision. Discuss both direction and magnitude of any potential bias | Discussion ('Strengths and limitations of the study' subheading) | RECORD 19.1: Discuss the implications of using data that were not created or collected to answer the specific research question(s). Include discussion of misclassification bias, unmeasured confounding, missing data, and changing eligibility over time, as they pertain to the study being reported. | Discussion ('Strengths and limitations of the study' subheading) |
| Interpretation | 20 | Give a cautious overall interpretation of results considering objectives, limitations, | Discussion ('Findings in context' subheading) | | |

| | | | | | |
|---|---|---|---|---|---|
| | | multiplicity of analyses, results from similar studies, and other relevant evidence | | | |
| Generalisability | 21 | Discuss the generalisability (external validity) of the study results | Discussion ('Strengths and limitations of the study' subheading) | | |
| **Other Information** | | | | | |
| Funding | 22 | Give the source of funding and the role of the funders for the present study and, if applicable, for the original study on which the present article is based | Funding | | |
| Accessibility of protocol, raw data, and programming code | | .. | | RECORD 22.1: Authors should provide information on how to access any supplemental information such as the study protocol, raw data, or programming code. | Link provided for Github repository containing codelists and code |

*Reference: Benchimol EI, Smeeth L, Guttmann A, Harron K, Moher D, Petersen I, Sørensen HT, von Elm E, Langan SM, the RECORD Working Committee. The REporting of studies Conducted using Observational Routinely-collected health Data (RECORD) Statement. *PLoS Medicine* 2015; in press.

*Checklist is protected under Creative Commons Attribution (CC BY) license.
